# Integration of a deep learning basal cell carcinoma detection and tumor mapping algorithm into the Mohs micrographic surgery workflow and effects on clinical staffing: a simulated, retrospective study

**DOI:** 10.1101/2023.05.13.23289947

**Authors:** Rachael Chacko, Matthew J. Davis, Joshua Levy, Matthew LeBoeuf

## Abstract

**Background:** Staffing shortages and inadequate healthcare access have driven the development of artificial intelligence (AI)-enabled tools in medicine. Accuracy of these algorithms has been extensively investigated, but research on downstream effects of AI integration into the clinical workflow is lacking.

**Objective:** We aim to analyze how integration of a basal cell carcinoma detection and tumor mapping algorithm in a Mohs micrographic surgery (MMS) unit may impact waiting times in the surgical pathology laboratory and on the floor.

**Methods:** Time spent on each task and slide, staff, and histotechnician waiting times were analyzed over a 20 day period in a MMS unit. A simulated AI workflow was created and the time differences between the real and simulated workflows were compared.

**Results:** Simulated addition of the algorithm led to improvements of 64% in slide waiting time (1:03:39 per case), 36% in staff waiting time (59:09 per case), and 25% in histotechnician waiting time (25:27 per case).

**Limitations:** A single MMS unit was analyzed and AI integration was performed retrospectively, rather than in real time.

**Conclusions:** AI integration results in significantly reduced slide, staff, and histotechnician waiting time, which enables increased productivity and a streamlined clinical workflow.

**Capsule summary:**

- The accuracy of artificial intelligence algorithms has been well established. This study addresses the impact of implementation of such an algorithm into a real-world clinical workflow.
- Results indicate the potential for increased efficiency and productivity with use of artificial intelligence in Mohs micrographic surgery.

## 1. Introduction

The introduction of artificial intelligence (AI) into medicine has long promised to improve healthcare delivery and the experiences of both patients and providers. The prevalence of AI applications within healthcare is increasing rapidly, as evidenced by the 343 FDA-approved AI-enabled medical devices currently in clinical practice (U.S. Food and Drug Administration, 2022). AI-enabled medical devices aim to provide diagnostic expertise where not otherwise available, improve clinicians’ diagnostic accuracy, and increase diagnostic efficiency while reducing fatiguability of redundant tasks (Feng et al., 2018; Feng et al., 2019; Puri et al., 2020). Despite the numerous studies on the effectiveness and utility of AI-based algorithms, research regarding implementation of these algorithms into the real-world clinical workflow is lacking.

Evolving over the last decade and hastened by the pandemic, nationwide staffing shortages have presented a major challenge to healthcare delivery (Chervoni-Knapp, 2022). An aging population with increased healthcare needs will continue to drive demand as highlighted by the United States Bureau of Labor Statistics’ estimate that over 195,000 additional registered nurses and 112,000 medical assistants will be necessary by 2031 (Bureau of Labor Statistics, 2022). In the current healthcare environment, utilization of AI in the real-world clinical setting has been limited by uncertainty from patients and providers as well as limited generalizability of algorithms beyond ideal situations created in a research setting (Young et al., 2021; Young et al., 2021; Yildirim et al., 2021). However, with improved understanding and study of the downstream effects of AI on process and staff efficiency, it may provide one solution to addressing the increased staffing demands of healthcare.

The highly specialized process of Mohs micrographic surgery (MMS) is especially suited for implementation of such algorithms to improve clinical workflow efficiency. Throughout each day, rate-limiting steps including tissue processing and histologic analysis can impact the efficiency of this procedure and result in increased patient and staff waiting time. AI provides an opportunity to further streamline this process and decrease this waiting time to improve patient experience (Supplementary Figure 1).

While numerous studies have shown the use of AI algorithms for diagnosis or analysis, investigation into the effects of AI on staff efficiency and clinical workflow are lacking. One of the main bottlenecks in the MMS workflow where AI integration can be useful is the time that slides spend waiting to be analyzed by the Mohs micrographic surgeon. We have developed a BCC detection algorithm to be used in the context of MMS that provides tissue grossing and inking recommendations, histologic BCC identification, and tumor mapping (Levy et al., 2022). Aimed at decreasing the rate-limiting steps of the iterative MMS process, the BCC detection algorithm was shown to accurately and rapidly identify and map tumor, hypothetically reducing or eliminating the need of the Mohs surgeon to perform histologic examination of the tissue margins. In this study, the BCC detection and tumor mapping algorithm is used in the context of a MMS unit to simulate how decreasing an important rate-limiting step through AI incorporation may affect clinical and laboratory staff efficiency.

## 2. Materials and Methods

The study included 108 consecutive MMS BCC cases performed over a 20-day period. To create a simulated MMS workflow that integrates the BCC detection algorithm, we defined three measures to evaluate algorithm integration. slide waiting time, staff waiting time, and histotechnician waiting time, depicted in Figure 1. *Slide waiting time* is defined as the time between when the histotechnician places the prepared slides next to the microscope and when the surgeon performs histologic analysis of the slides and subsequent tumor mapping. *Staff waiting time* is the time between when the histotechnician places the prepared slides next to the microscope and when the nursing staff member begins preparation for the next step, which is either an additional stage or defect repair. *Histotechnician waiting time* is the time between when the histotechnician places the prepared slides next to the microscope and when the surgeon delivers tissue from the next stage in cases in which additional tumor removal is required. On the days of surgery, slide, staff, and histotechnician waiting times were measured for each of the 108 cases. To generate simulated waiting times, the BCC detection algorithm was implemented at the immediate time the slides were placed next to the microscope.

**Figure 1.**
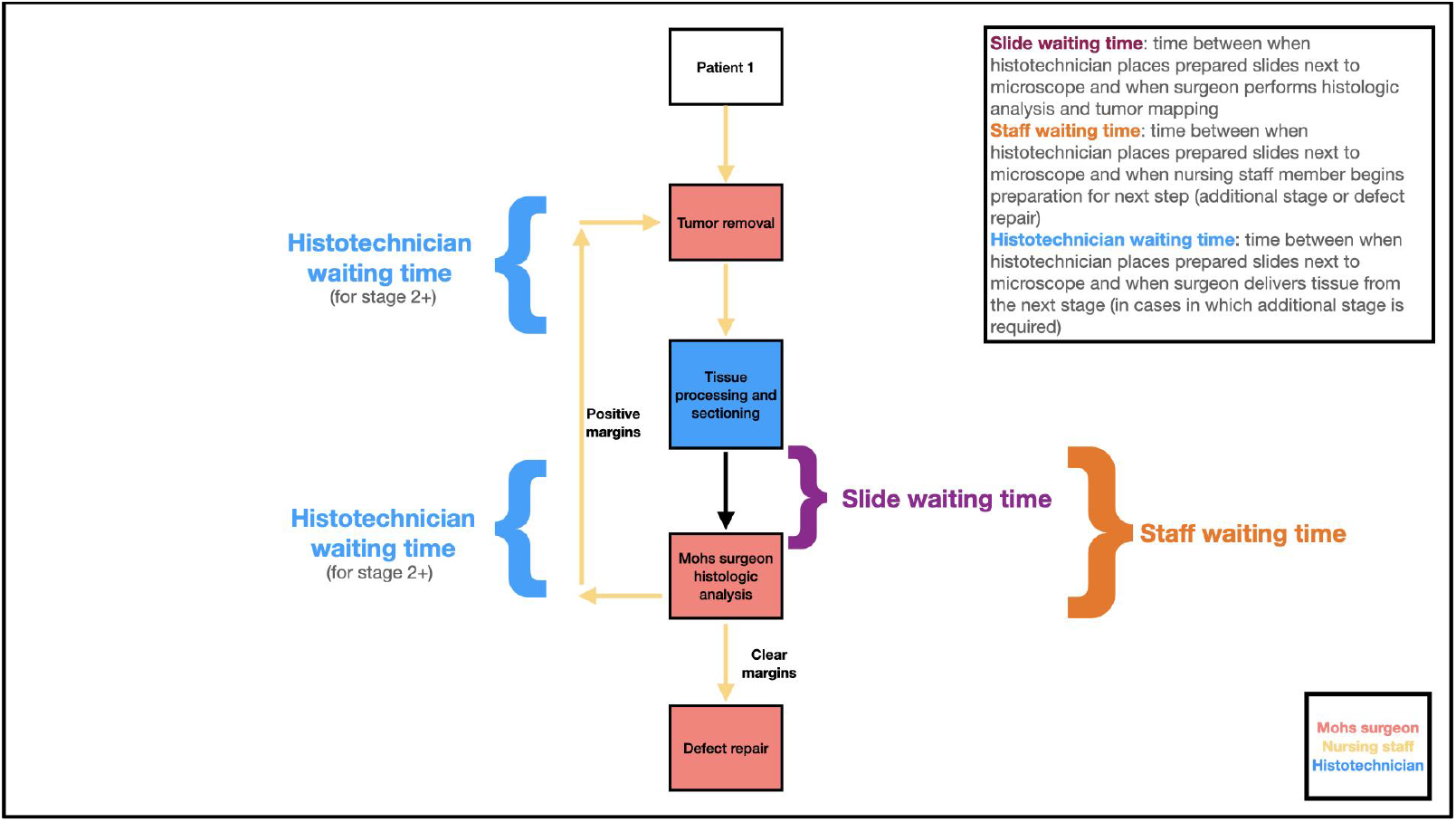
Definitions and depiction of slide, staff, and histotechnician waiting times.

To simulate algorithm integration as accurately as possible, we measured slide scanning time, algorithm processing time, and algorithm runtime (Supplementary Figure 2). The sum of these three components represents the simulated slide waiting time. Sixty-five cases reflecting the normal distribution of BCC histologic subtypes in MMS clinical practice were selected to undergo slide scanning at 20X using an Aperio AT2 to generate whole slide images (WSI) in the SVS format with 8-bit color channels. We recorded the scanning time per slide and summed per case to determine total slide scanning time. WSI were assessed using the BCC detection and tumor mapping algorithm, which has been previously described, and algorithm processing time and runtime were measured. All cases were deidentified for the comparison of algorithm-versus surgeon-generated tumor maps. Concordance between the tumor maps was established based on a retrospective review and subjective interpretation of visual agreement (yes/no). The proportion of times the two maps agreed was recorded as a measure of concordance and 95% confidence intervals were obtained using a normal approximation of the binomial probabilities. No clinical decisions were made using algorithm output.

Lastly, to generate simulated staff and histotechnician waiting times for each case, we then took the difference between the actual and simulated slide waiting times, which represents the time saved or lost with algorithm integration. This amount of time was then subtracted or added from the downstream staff and histotechnician waiting times.

## 3. Results

Removal of skin cancer with MMS relies on a clinical workflow involving nursing staff, the Mohs micrographic surgeon, and histotechnicians in the on-site pathology laboratory. Staffing numbers and operating room space can affect the length of time required to remove the tumor with iterative real-time histologic margin analysis and subsequent repair of the resulting surgical defect. Over the 20-day study period, the numbers of nursing staff (4), histotechnicians (2), and operating rooms (5) were held constant. Additional variables hypothesized to affect staff and histotechnician waiting times included number of tumor removal stages per day and repair complexity.

Critical to the introduction of AI into clinical practice is to ensure the algorithm performs at or above the level of the human expert. To test this, a cohort of 65 BCC cases reflecting the normal distribution of BCC histologic subtypes in MMS clinical practice were scanned and analyzed by the BCC detection algorithm (Table I). The algorithm-generated tumor maps were compared to the hand-drawn tumor maps created by the Mohs surgeon at the time of surgery (Figure 2). The BCC detection algorithm identified the tumor and appropriate location of tumor in 94% (95% CI: 88%-99%) of surgical cases.

**Table I.** Selected BCC case histologic subtypes and algorithm accuracy. All cases were deidentified for the comparison of algorithm-versus surgeon-generated tumor maps. Concordance between the tumor maps was established based on a retrospective review and subjective interpretation of visual agreement (yes/no). The proportion of times the two maps agreed was recorded as a measure of concordance and 95% confidence intervals were obtained using a normal approximation of the binomial probabilities. Non-parametric bootstrapping was used to obtain 95% confidence intervals for the timing estimates.

|  |  |
| --- | --- |
| **Number of Cases** | 65 |
| **BCC Histologic Subtype** |  |
| --Nodular | 37 |
| --Superficial | 14 |
| --Infiltrative | 15 |
| --Micronodular | 5 |
| --No tumor | 16 |
| **Accuracy (CI)** | 0.94 (0.88-0.99) |
| **Median slide scanning time (s)** | 180 |
| **Median algorithm processing and execution time (s)** | 69.21 |

**Figure 2.**
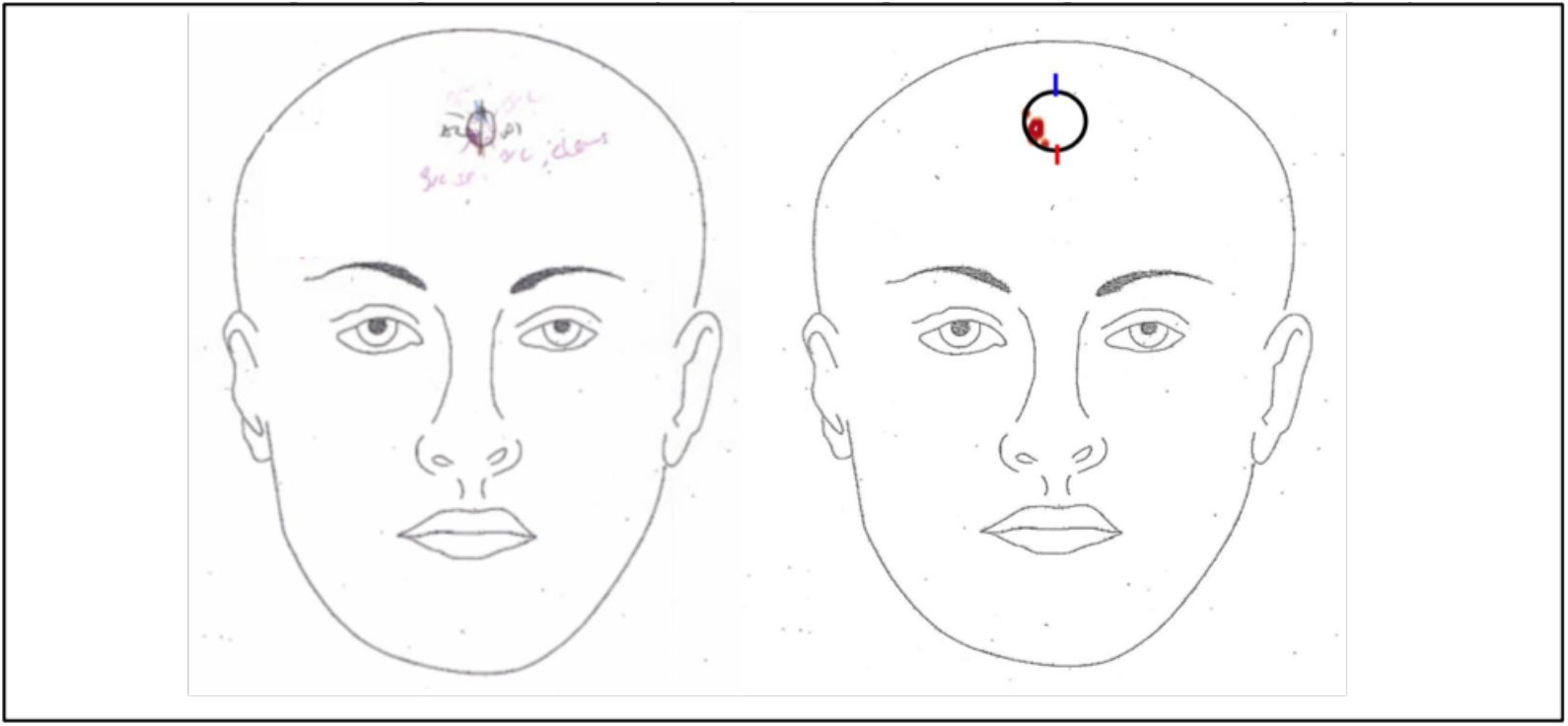
Surgeon-generated (left) vs. algorithm-generated (right) tumor maps.

To determine how the BCC detection and tumor mapping algorithm might affect the MMS workflow, a simulated workflow was created in which histologic assessment was performed by the BCC detection algorithm. The amount of time required to scan the selected 65 BCC cases and generate output from the algorithm was measured for each case (Supplementary Table I). The median slide scanning time and algorithm computation time per case were 180 and 69.21 seconds, respectively. Simulated waiting times were adjusted by adding or subtracting the slide scanning/algorithm output time from the actual slide waiting time for each of the 108 BCC cases. On average, 65% of slide waiting time across the 20-day period was saved with the simulated addition of the algorithm, which translated to an average of 1:05:22 per case. As a result, 36% of staff waiting time (1:00:09 per case) and 27% of histotechnician waiting time (27:00 per case) was saved (Table II).

**Table II.** BCC algorithm implementation effects on slide, staff, and histotechnician waiting time. Wilcoxon signed-rank tests were used to assess whether the differences between actual and simulated waiting times were significant.

|  |  |
| --- | --- |
| **Average slide waiting time saved (h:mm:ss)** | 1:05:22 (p<0.001, W=37) |
| **Average % slide waiting time saved** | 65.26% |
| **Average staff waiting time saved (h:mm:ss)** | 1:00:09 (p<0.001, W=41) |
| **Average % staff waiting time saved** | 36.49% |
| **Average histotechnician waiting time saved (h:mm:ss)** | 0:27:00 (p<0.001, W=70) |
| **Average % histotechnician waiting time saved** | 26.61% |

## 4. Discussion

Increasing demands for healthcare combined with nationwide medical staffing shortages and decreasing reimbursement rates pose a problem for healthcare delivery to which AI may offer a solution. Here we demonstrate that simulated use of a BCC detection and tumor mapping algorithm within the MMS workflow can reduce significant slide, staff, and histotechnician waiting time. By replacing the requirement of the surgeon to perform the real-time histologic examination and tumor mapping, subsequent steps in the MMS process can be carried out in a more efficient manner. Notably, the nursing staff can act at an earlier timepoint to prepare both the operating room and patients for either additional tumor removal or defect reconstruction. The surgeon can move between operating rooms without the normal waiting time required for patient and staff transition. This results in more efficient tumor removal and subsequent defect repair. Histotechnicians receive tissue in the lab at an earlier time point than they would otherwise, decreasing the overall amount of histotechnician time required. Together, algorithm implementation in the simulated scenario increased the parallelization of an otherwise serial process. This demonstrated that integration of the BCC detection algorithm for histologic analysis resulted in a significantly decreased amount of staff and histotechnician waiting time through real-time immediate histologic analysis and tumor map output.

In identification of processes in healthcare that would benefit from the introduction of AI, additional factors outside of pure diagnostic accuracy need to be considered. Processes that are repetitive or highly iterative and where multiple or numerous individuals rely on decision making from a single individual may be ideal. Perspectives and feedback from all members of the clinical care team that might be affected by AI integration should be taken into account. Design of algorithms to be used in settings with staffing shortages or where there is a mismatch in diagnostic information and staff action should be considered. Detailed analysis of clinical workflows including all members of the team may highlight significant bottlenecks and inefficiencies that could be solved by the introduction of AI-enabled tools.

This simulated workflow thus far has only been run retrospectively and at a single site, and actual clinical implementation may slightly alter outcomes. The cost of implementation and complexity of such a robust automated system is significant. AI integration in the MMS clinical workflow requires a slide scanner, elevated levels of computing power, and reliable access to high-speed internet. Not only are these variables expensive, but as with any technology-dependent system, inferior performance of any of these components may significantly impact real-life results. A median algorithm runtime of 69.21s was used to approximate the runtime for each case. Future studies could involve using each individual case’s runtime for waiting time calculations, though we do not believe this change is likely to make a clinically significant difference. Next steps include slide collection and waiting time analysis at multiple MMS units to investigate the extent to which AI integration can impact MMS units with varying numbers of Mohs surgeons, nursing staff, histotechnicians, and available operating rooms. Analysis of how the complexity of defect repairs and the number of stages of tumor removal per day affect slide waiting time is also necessary to evaluate the generalizability of this simulated workflow. As additional tumor detection algorithms are developed, similar workflow analysis may be done to assess the generalizability of this study to other tumor types.

While medicine faces many healthcare infrastructure challenges including adequately staffing our hospitals and clinics, appropriate and thoughtful introduction of digital technology offers one solution. Design and testing validation of AI solutions by end users, including providers and medical staff, offers an opportunity to correctly identify important clinical variables that need to be controlled for and the most relevant bottlenecks in the healthcare delivery process. Early analysis of both positive and negative neighborhood effects, including cost and resource requirements, will help identify clinical settings most likely to benefit from AI implementation. This study addresses and controls for multiple clinical variables and highlights how removing a rate-limiting step in the delivery of surgical and pathologic care results in significantly increased staff efficiency.

## Data Availability

Due to data privacy restrictions, subsets of the data produced in the present study may be made available upon reasonable request to the authors.

## Supplementary Figures and Tables

**Supplementary Figure 1.**
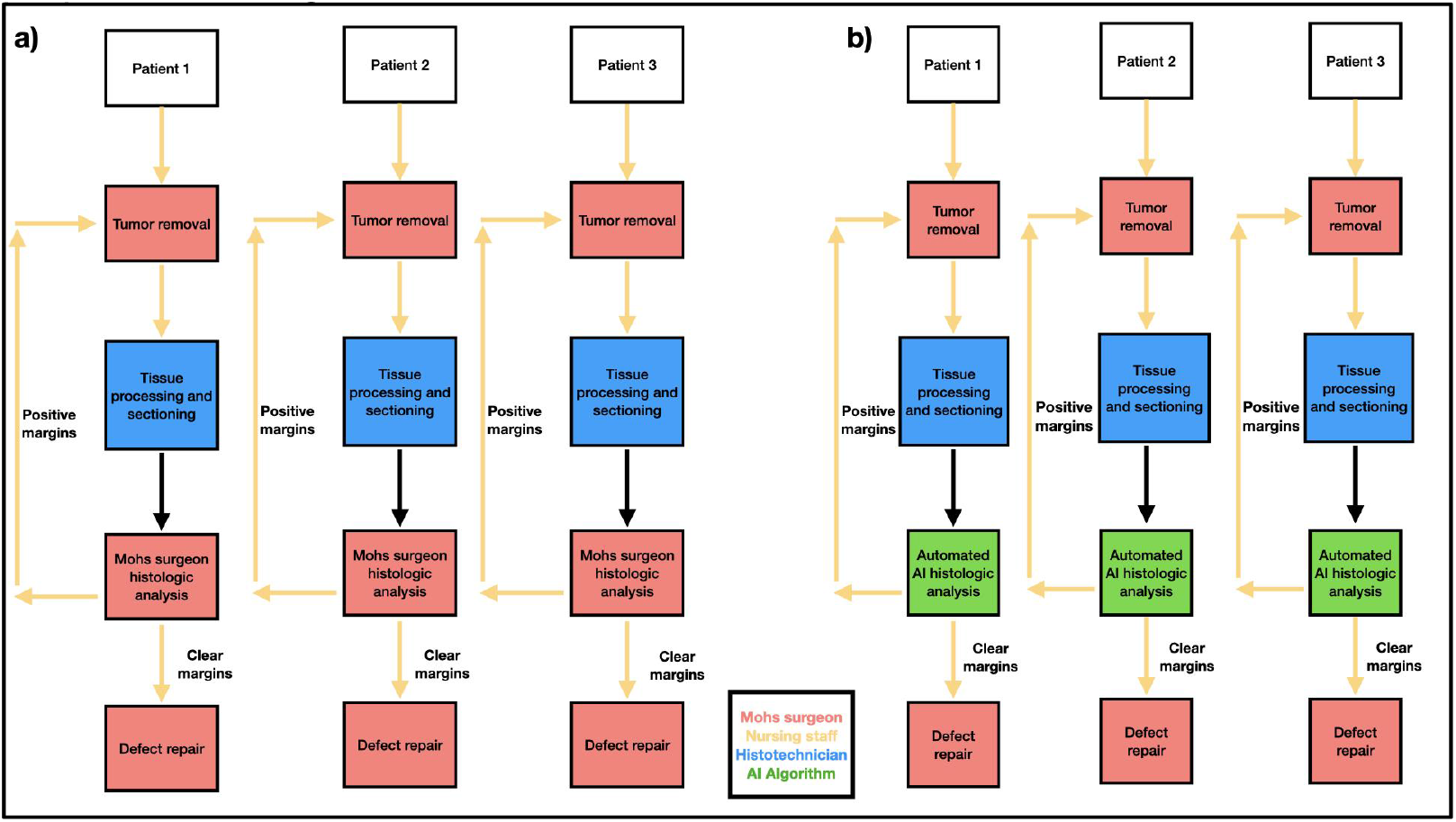
a) Current MMS clinical workflow. b) MMS clinical workflow with proposed AI integration.

**Supplementary Figure 2.**
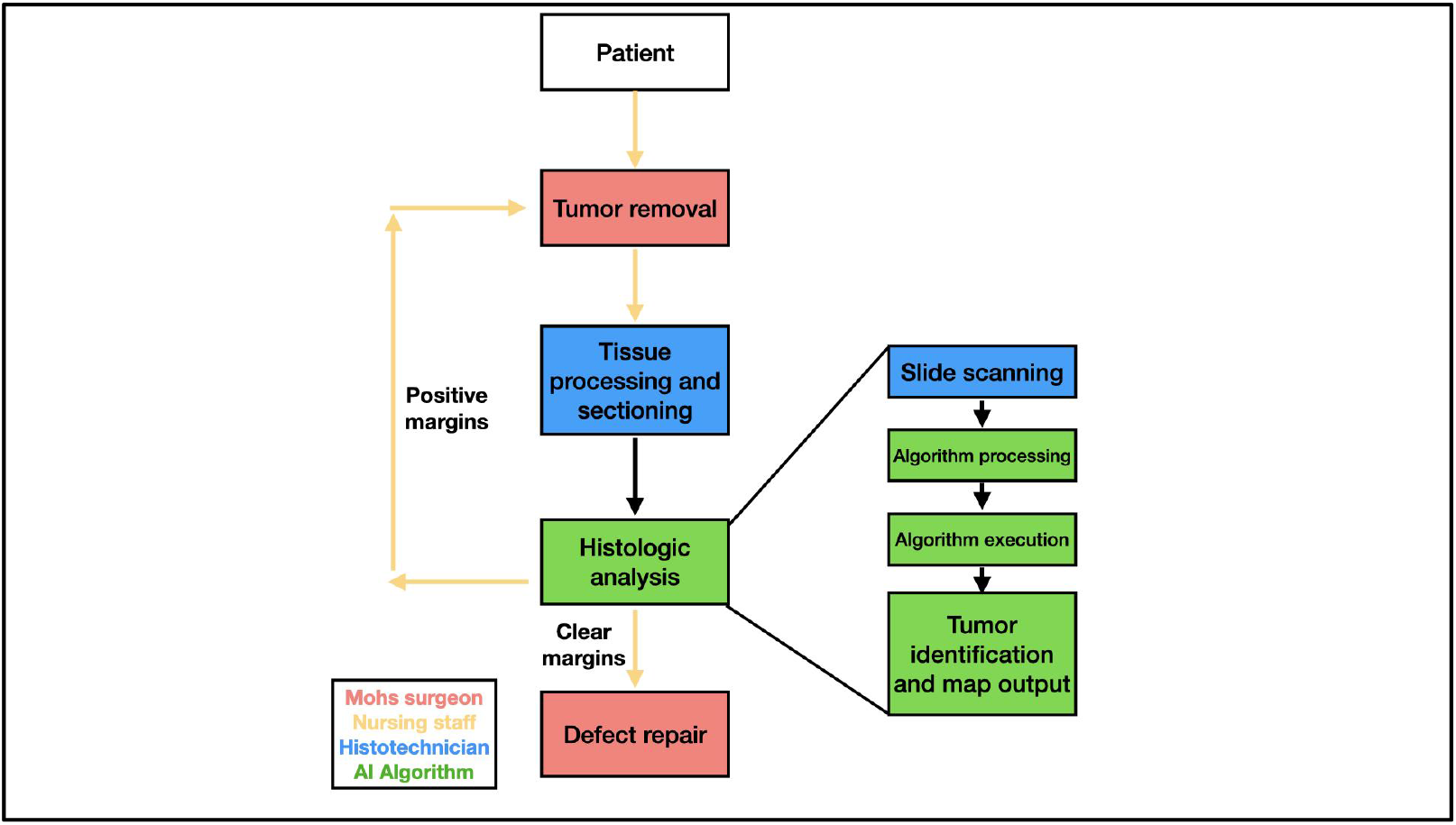
AI-augmented MMS workflow. *Algorithm processing time* is the time required for WSI upload into the algorithm platform. *Algorithm execution/runtime* is the time required for generation of the tumor map and clinical decision output.

**Supplementary Table I.**
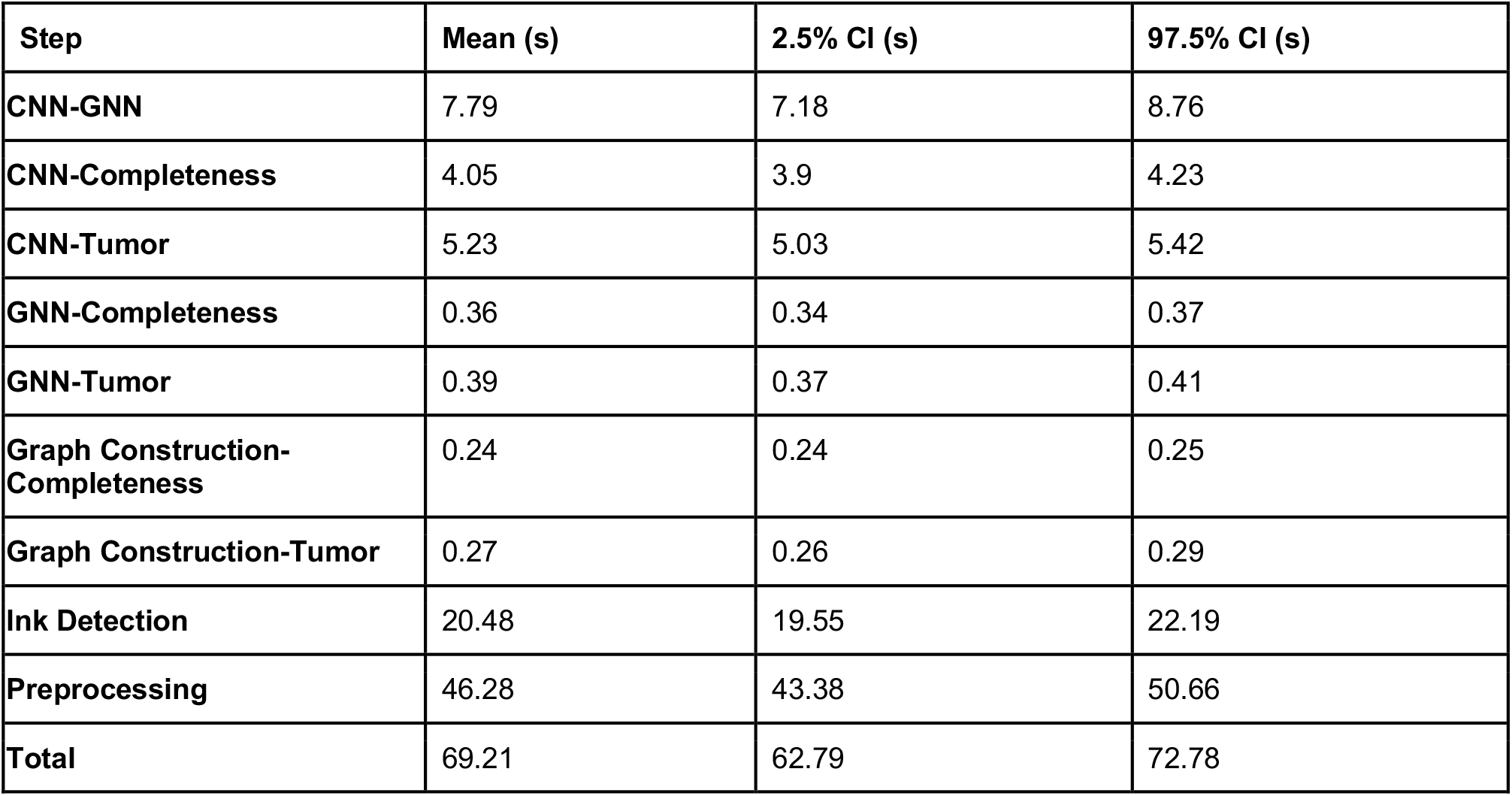
Mean and 95% Confidence Interval for time elapsed for each step in the histologic analysis process.

## Notes

**Funding sources:** JL is supported by NIH grants R24GM141194, P20GM104416 and P20GM130454.

**Conflicts of Interest:** None declared.

### Competing Interest Statement

The authors have declared no competing interest.

### Funding Statement

JL is supported by NIH grants R24GM141194, P20GM104416 and P20GM130454.

### Author Declarations

Human Research Protection Program of Dartmouth Health gave ethical approval for this work.

